# High-throughput assay confirmation of a T-cell receptor pre-mRNA fragment as a blood-based inflammatory breast cancer biomarker

**DOI:** 10.1101/2025.08.19.25333925

**Authors:** Elizabeth A. Ferrick-Kiddie, Sriya Dommaraju, Jun Yao, Christopher W. Smith, Xiaoping Wang, Wendy A. Woodward, The MDACC Inflammatory Breast Cancer Team, Naoto T. Ueno, Savitri Krishnamurthy, Alan M. Lambowitz

## Abstract

Previous TGIRT-seq analysis of RNAs in Inflammatory Breast Cancer (IBC) patient tumors, peripheral blood mononuclear cells (PBMCs) and plasma identified a short T-cell receptor mRNA fragment (*TRBJ1-6*) as a potential IBC biomarker that was detected in plasma samples from IBC patients but not patients with non-inflammatory breast cancer or healthy donors. Here, we traced the origin of this *TRBJ1-6* RNA fragment to IBC patient PBMCs and used a high-throughput RT-PCR/Cas12a assay with larger numbers of samples to confirm its prevalence in IBC patient PBMCs. Detection of this RNA was enhanced by T4 polynucleotide kinase treatment, indicating the presence of a 2’,3’-cyclic phosphate. Analysis of previous TGIRT-seq datasets revealed gene expression differences in IBC patient PBMCs that could contribute to *TRBJ1-6* RNA prevalence in IBC patient PBMCs and plasma. Our results support the identification of the *TRBJ1-6* RNA fragment as a novel, readily detectable blood-based RNA biomarker derived from IBC-patient immune cells, addressing a major unmet need for diagnosing IBC.

**Teaser:** A novel type of blood-based biomarker for diagnosis of Inflammatory Breast Cancer

## Introduction

Inflammatory Breast Cancer (IBC) is a rare and aggressive breast cancer variant that has a disproportionately high fatality rate and is diagnosed minimally at stage III with ∼30% of IBC patients diagnosed at stage IV (*1–3*). IBC also has a substantial initial misdiagnosis rate that delays treatment due to its clinical presentation with breast skin discoloration and edema that are mistaken for breast infections, its frequent lack of a palpable tumor lump, its diffuse pattern of spread that can be missed on mammograms, and the lack of biomarkers that discriminate it from other breast cancers types (*1*, *2*, *4–6*). Earlier diagnosis that could improve treatment decisions and outcomes for IBC patients would be greatly helped by the discovery of blood-based biomarkers. In a previous study, we used Thermostable Group II Intron Reverse Transcriptase sequencing (TGIRT-seq) to comprehensively profile protein-coding and non-coding RNAs in Formalin-Fixed Paraffin Embedded (FFPE) tumors, Peripheral Blood Mononuclear Cells (PBMCs), and plasma samples from IBC patients, patients with non-inflammatory breast cancer (non-IBC) subtypes, and healthy donors and identified differentially expressed RNAs in all three sample types (*7*). A promising, potential IBC biomarker identified by TGIRT-seq in 10 of 10 IBC patient plasma samples was an RNA fragment that originated from a T Cell Receptor Beta Joining 1-6 (*TRBJ1-6*) gene segment derived from a larger T Cell Receptor Beta gene (TCRβ) transcript. This *TRBJ1-6* RNA was short (20-23 nt) and comprised of a 5’ exon sequence followed by a short intron sequence with a specific 3’ terminus, suggesting that it arose from a messenger RNA precursor (pre-mRNA) (*7*).

Here, we found that this *TRBJ1-6* pre-mRNA fragment could also be detected by TGIRT-seq in IBC patient PBMCs and validated its higher differential expression levels in larger PBMC sample sets by using a high-throughput RT-PCR/Cas12a method related to those used for detection of miRNAs. Further analysis indicated that the *TRBJ1-6* RNA fragment is stabilized at least in part by a 2’,3’-cyclic phosphate end modification that could impede its detection by other RNA-seq library preparation methods. Our findings address a major unmet need for the diagnosis of IBC and support the importance of blood-based biomarkers corresponding to RNA fragments that are stabilized by a 2’,3’-cyclic phosphate.

## Results

### Origin of the *TRBJ1-6* biomarker

The *TRBJ1-6* RNA fragment identified by comprehensive TGIRT-seq as a putative IBC biomarker was a 20-23 nt RNA fragment that spans an exon-intron junction located 63 nucleotides downstream of the beginning of the *TRBJ1-6* exon segment and has slightly variant 5’ ends and a specific 3’ end (*7*). These findings suggested that it could be derived from either a VDJ recombined or unrecombined TCRβ RNA transcript. Our previous TGIRT-seq analysis identified the *TRBJ1-6* RNA fragment as differentially enriched in IBC patient plasma samples, but Integrated Genomics Viewer (IGV) alignments for multiple patient samples also showed discrete RNA peaks at a similar position in TGIRT-seq datasets of IBC patient PBMC and tumor samples (Figure 1A; (*7*)). When the PBMC total RNA TGIRT-seq libraries were bioinformatically filtered to keep only short transcripts (≤30 nt), the discrete *TRBJ1-6* fragment was seen to be significantly upregulated in 8 of 10 IBC patient PBMC samples and not detected in non-IBC patient or healthy donor PBMCs, indicating that it was generated in IBC patient PBMCs prior to its appearance and enrichment in plasma (Figure 1A and B). Although not detected in two IBC patient PBMCs samples, the same *TRBJ1-6* RNA fragment was detected in plasma samples from the same two IBC patients (*7*), suggesting that failure to detect it in some PBMCs reflects lower read depth relative to other RNAs in highly activated IBC patient PBMC total-RNA libraries (*7*). Peaks corresponding to *TRBJ1-2, TRBJ1-3, TRBJ1-4, and TRBJ1-5* RNA fragments were also observed in some of the size-filtered PBMC datasets but were not as consistently enriched in IBC patient PBMCs as the *TRBJ1-6* RNA fragments (Figure 1A).

**Figure 1.**
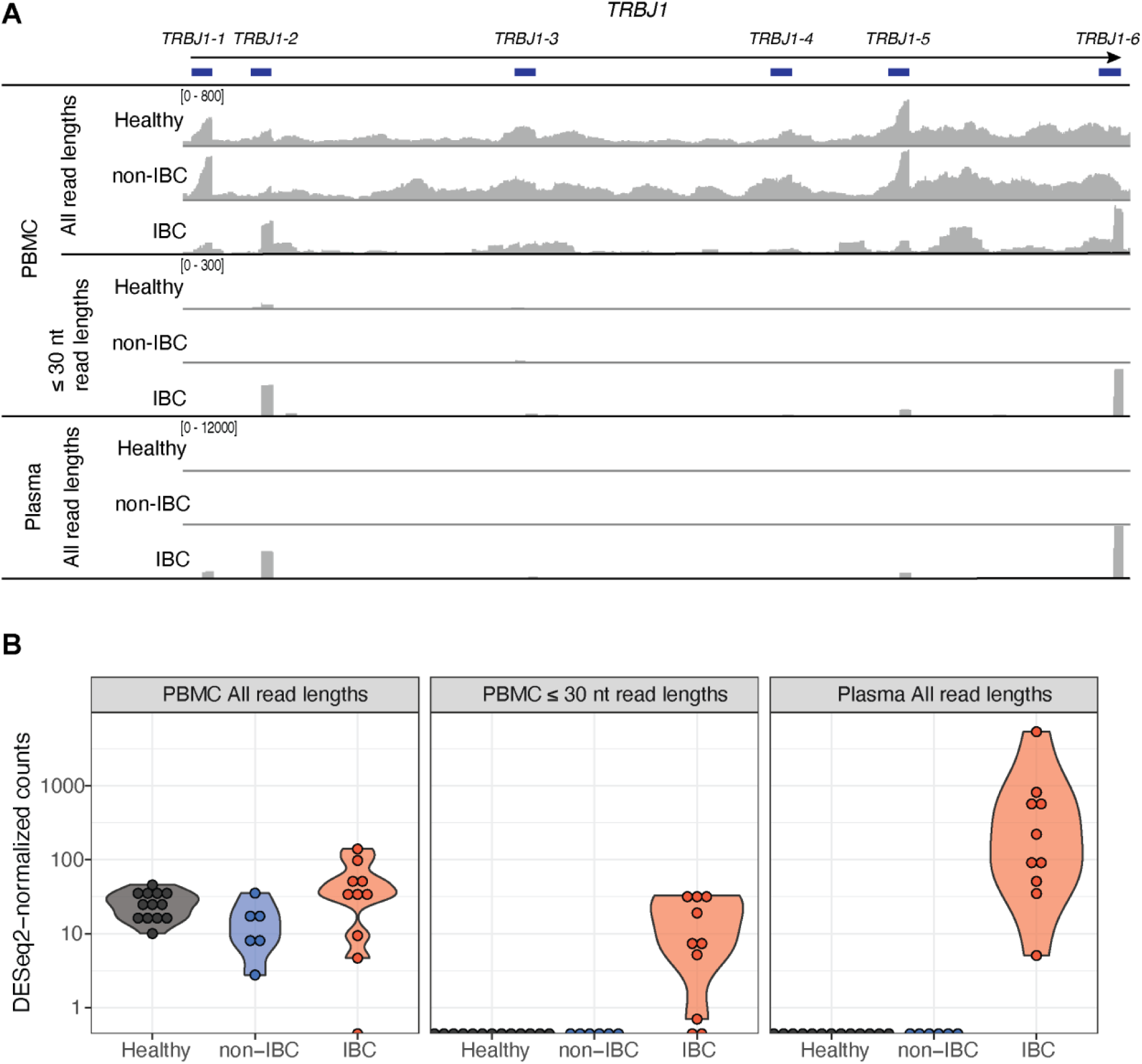
Short *TRBJ1* RNA fragments are enriched in IBC patient PBMCs and plasma. **(A)** Integrated Genomics Viewer (IGV) plots of TGIRT-seq reads of RNAs mapped to the portion of the *TRBJ1* genomic locus encompassing *TRBJ1-1* through *TRBJ1-6* exon gene segments (blue bars) in TGIRT-seq combined datasets for 10 IBC patients, 6 non-IBC patients, and 13 healthy donor PBMCs and plasma samples (*7*). y-axis read depth ranges in combined datasets for each comparison are indicated in brackets at the upper left of each plot. The top and bottom plots are based on paired-end reads of all lengths for PBMCs and plasma and the middle plot is based on paired-end read lengths ≤30 nt for PBMCs. **(B)** Violin plots of *TRBJ1-6* reads in IBC patient PBMCs and plasma compared to non-IBC patient and healthy donor PBMCs and plasma. *TRBJ1-6* reads were mapped to the genomic reference and read counts were normalized using DESeq2 as done in Wylie et al. (*7*). TGIRT-seq datasets for PBMCs were also bioinformatically filtered to include only paired-end protein-coding RNAs that were ≤30 nt, followed by DESeq2 differential gene expression analysis. Reads for short *TRBJ1-6* RNAs were significantly enriched in plasma from IBC patients compared to plasma from healthy donors (p_adj_ = 4.04E-13) and non-IBC patients (p_adj_ = 1.43E-8) and in PBMCs filtered to ≤30 nt read lengths from IBC patients compared to healthy donors (p_adj_ = 2.86E-4) and non-IBC patients (p_adj_ = 9.59E-5).

### Design of a high throughput RT-PCR/Cas12a assay for *TRBJ1-6* RNA biomarker

To support validation of the *TRBJ1-6* biomarker in PBMC RNAs, we sought a high-throughput method for testing larger numbers of samples (Figure 2). Because the *TRBJ1-6* mRNA fragment is small and miRNA-sized, we focused on miRNA-detection methods. A common method for detection of miRNAs involves the addition of a poly(A) tail by poly(A) polymerase followed by oligo(dT)-primed reverse transcription for first strand cDNA synthesis and PCR using a target specific forward primer and a reverse primer corresponding to a universal sequence incorporated in the oligo(dT) reverse transcriptase (RT) primer (Figure 2 and Table S1). For our assay, we combined two previously described methods: Niu *et al.* (*8*) in which poly(A) tailing is done together with reverse transcription, and Zhong *et al.* (*9*), which uses the artificially added poly(A) tail to create a Protospacer Adjacent Motif (PAM) sequence in the resulting PCR amplicon for detection by Cas12a in complex with a target specific crRNA, a method referred to as PCDetection (PolyA-CRISPR/Cas12a-based miRNA detection without PAM restriction) (Figure 2). The PCDetection method is a continuation of the exciting new field of CRISPR/Cas12a-based diagnostics, where a specifically generated PCR target is used to prime the initial activation of the Cas12a enzyme in complex with a target-specific CRISPR RNA (crRNA); once the Cas12a/crRNA complex is activated it can exhibit trans-cleavage activity on a fluorescent reporter probe (*9–15*). In our system, the target-specific crRNA was designed to recognize the precise 3’ end of the *TBRJ1-6* RNA based on the location of the artificially added PAM, but retains some flexibility at its 5’ end, as we observed that the *TRBJ1-6* RNA fragment had a specific 3’ end in all previously analyzed IBC patient PBMC and plasma samples but could vary by up to three nucleotides at the 5’ end (Table S1) (*7*).

**Figure 2.**
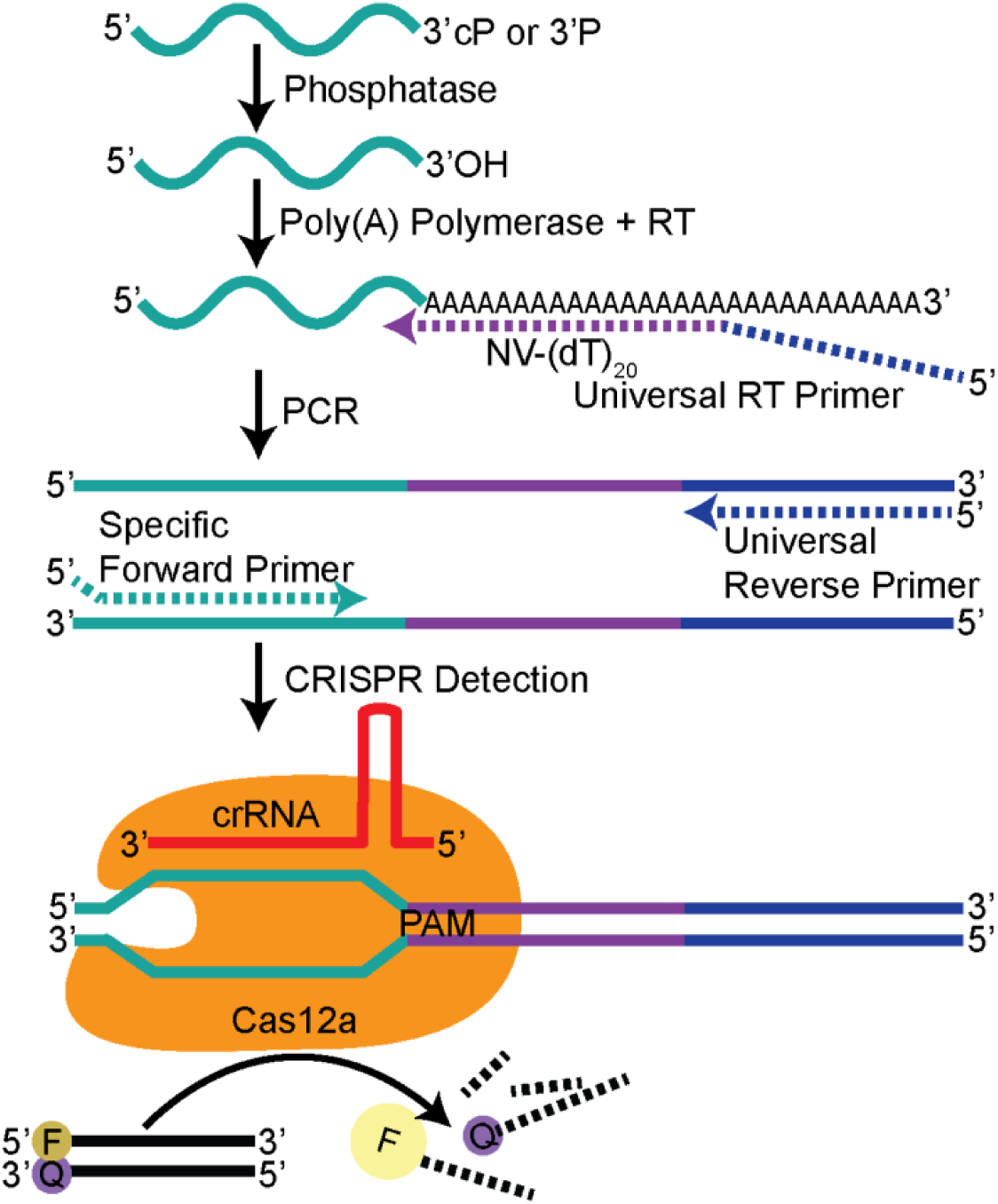
Schematic of high-throughput RT-PCR/Cas12a assay. RNA isolated from PBMCs from IBC and non-IBC patients and healthy donors was subjected to a phosphatase treatment, either PNK or CIP in an initial test assay (Figure 3) and PNK in subsequent final assays to remove 2’,3’-cyclic phosphates (Figure 4). Next, a poly(A) tail was added to serve as the complementary sequence for a reverse transcription (RT) primer consisting of an anchored oligo(dT)_20_ primer (NV-(dT)_20_, purple, NV signifies degenerate bases in the primer: N signifies any deoxynucleotide, dN, and V signifies either dG, dA, or dC) preceded by a 5’ universal reverse primer sequence (blue) that does not have significant similarity to any GenBank sequence (Table S1) (*8*, *56*). Following PCR with a gene-specific forward primer and the universal reverse primer, a Cas12a enzyme bound to a target-sequence specific CRISPR RNA (crRNA) was hybridized with the PCR amplicon, activating a *trans*-cleavage activity of Cas12a to shred a dsDNA fluorescence reporter (*13*). The resulting fluorescence increase was measured over a 180-min time course. Sequences of RT primers, PCR primers, and crRNAs are listed in Table S1.

Different endonucleases and exonucleases that exist in cells and plasma can leave different 3’ end modifications, some of which can impede detection by external validation assays. As pre-mRNA fragments may have a 3’-phosphate instead of a 3’-hydroxyl (3’-OH) group, due to endo- or exonucleolytic cleavage (*16*), we tested the addition of a phosphatase step to ensure the presence of free 3’-OH ends needed for the addition of a poly(A) tail by poly(A) polymerase (*17*). The two phosphatases tested were Calf Intestinal Phosphatase (CIP), which can efficiently remove 3’-phosphates (3’P) but not 2’,3’-cyclic phosphates (2’,3’cP) and T4 Polynucleotide Kinase (PNK), which can remove both 3’P and 2’,3’cP, leaving a 3’-OH (*16*). PNK is commonly used to enrich RNAs whose 3’ ends are otherwise blocked by 2’,3’cP RNAs in RNA sequencing methods (*16*, *18–26*), including in the previous TGIRT-seq analysis, which identified *TRBJ1* RNA fragments as potential IBC biomarkers (*7*).

An initial test of the RT-PCR/Cas12a method using small subsets of IBC patient, non-IBC patient, and healthy donor PBMC samples found that PNK treatment of two of the samples resulted in substantially increased detection of the *TRBJ1-6* RNA, to a greater extent than CIP treatment (Figure 3A and B), indicating the presence of a 2’,3’cP. In a third IBC patient PBMC sample, PNK was only slightly better than CIP in increasing detection of the *TRBJ1-6* RNA (Figure 3A and B), suggesting variations in the level 2’,3’cP-RNA that could impact detection in different patient PBMC samples (Figure 3A and B). Our tested PBMC subset also demonstrated that the newly developed RT-PCR/Cas12 assay worked as expected, with minimal background detection in controls that omitted poly(A)-tail addition, indicating no activation by full length *TRBJ1-6*-containing mRNAs, or omitting reverse transcription, indicating no activation by possible trace amounts of DNA (Figure 3A and Figure S1). PCR cycles were optimized individually for each target RNA in order to keep each assay below fluorescence saturation and fluorophore photobleaching (Figure 3A). mir-223-3p was chosen as a control that does not contain a 3’P and thus could be detected with or without PNK or CIP treatment (Figure 3A and B). Moving forward, we used PNK as the optimal phosphatase for the high-throughput assay.

**Figure 3.**
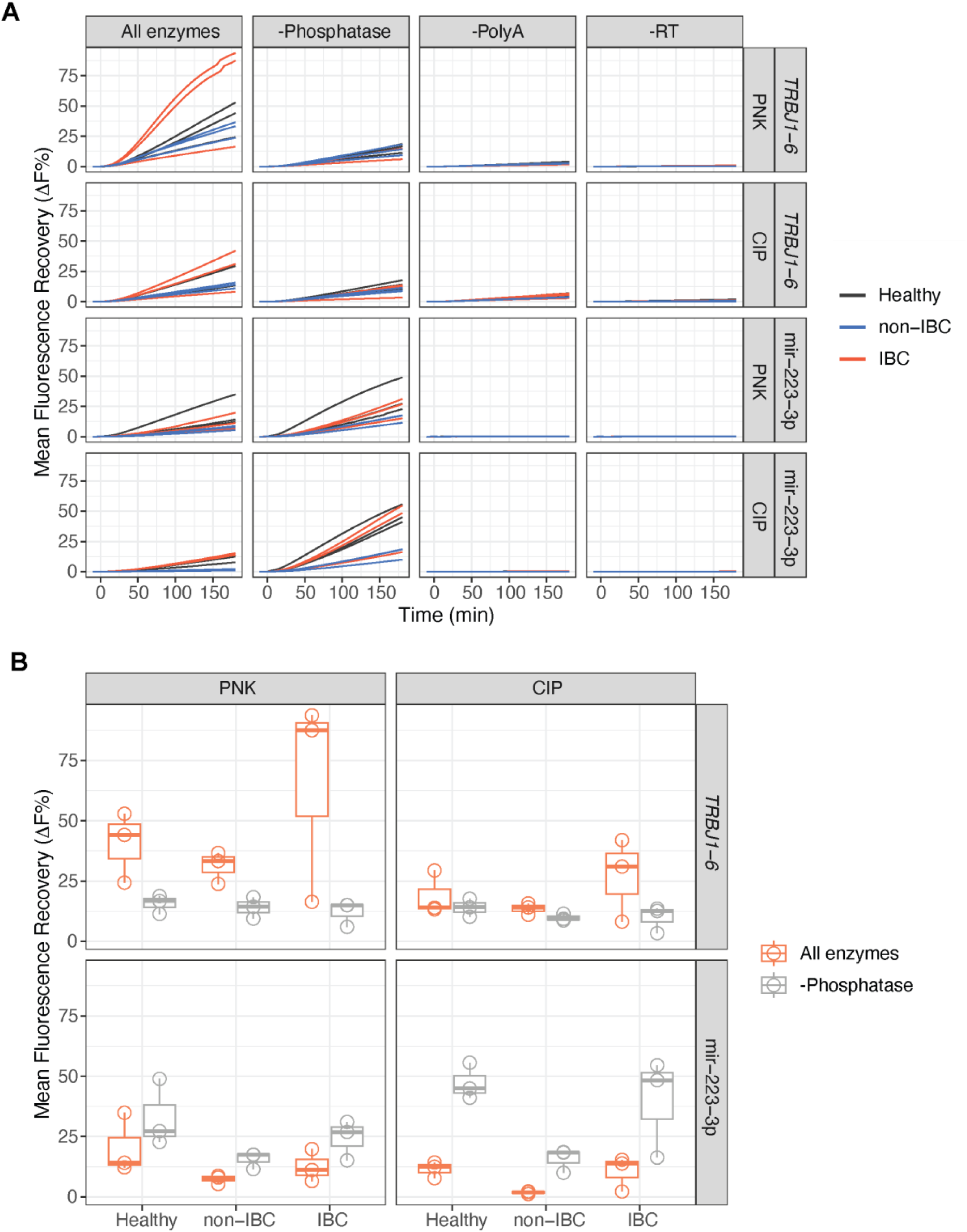
High-throughput assay for detection of *TRBJ1-6* RNA fragments and miRNA mir-223-3p in PBMCs and dependence on PNK treatment for detection of *TRBJ1-6* RNAs. **(A)** Time courses of RT-PCR/Cas12a assay detection for *TRBJ1-6* and mir-223-3p, with either PNK or CIP as the phosphatase. Control reactions were done without phosphatase (-Phosphatase, either PNK or CIP), Poly(A) polymerase (-Poly(A)), or reverse transcriptase (-RT). **(B)** Box plots comparing the detection of *TRBJ1-6* and mir-223-3p target RNAs that had been treated with or without PNK or CIP phosphatase, at the 180-min time point. The average value of two technical replicates for each of the three patient PBMC sample was used to calculate Mean Fluorescence Recovery (ΔF%).

### High-throughput validation by RT-PCR/Cas12a assay of the *TRBJ1-6* RNA biomarker in larger numbers of clinical PBMC samples

Having optimized the reactions, we next broadened the RT-PCR/Cas12a assays to include a larger number of PBMC samples: 32 IBC patients, 30 non-IBC patients, and 29 healthy donors, none of which had been analyzed previously by TGIRT-seq (Figure 4). As IBC is diagnosed at a minimum of stage III, in order to ensure the most relevant sample comparisons, all non-IBC patients used for comparisons were also diagnosed as stage III (Table S2). These assays confirmed that *TRBJ1-6* is a highly significant and sensitive biomarker for diagnosing IBC in PBMCs, compared to both healthy donor (adjusted p-value (p_adj_) = 1.15E-10) and non-IBC patient PBMCs (p_adj_ = 4.79E-10) (Figure 4A and B). Receiver Operating Curve (ROC) analysis for IBC versus combined healthy and non-IBC had an Area Under the Curve (AUC) of 0.979 (Figure 4C). In contrast, although the previous TGIRT-seq analysis of a limited number of IBC patient PBMC samples predicted that mir-223-3p might also be a diagnostic biomarker of IBC in PBMCs, mir-223-3p was elevated above the healthy and non-IBC detection in only a few IBC PBMC samples (Figure 4A and B). Separate comparisons between IBC patient and healthy donor or non-IBC patient PBMCs demonstrated the same trends as the combined healthy donor and non-IBC patient comparison (Figure 4C). Although we chose the time points with the maximum AUC to display in Figure 4, the AUC values across all time points of the RT-PCR/Cas12a assay were remarkably consistent (Figure S2). Although differences in *TRBJ1-6* levels in IBC patient and non-IBC patient and healthy donor PBMCs were highly significant, there was some overlap in *TRBJ1-6* levels between healthy donor and non-IBC patients and IBC patients with lower levels of *TRBJ1-6* RNA (Figure 4B). Assessing the nature and cause of these overlaps in PBMC samples will require comprehensive TGIRT-seq analysis of PBMC RNAs in these patient subsets.

**Figure 4.**
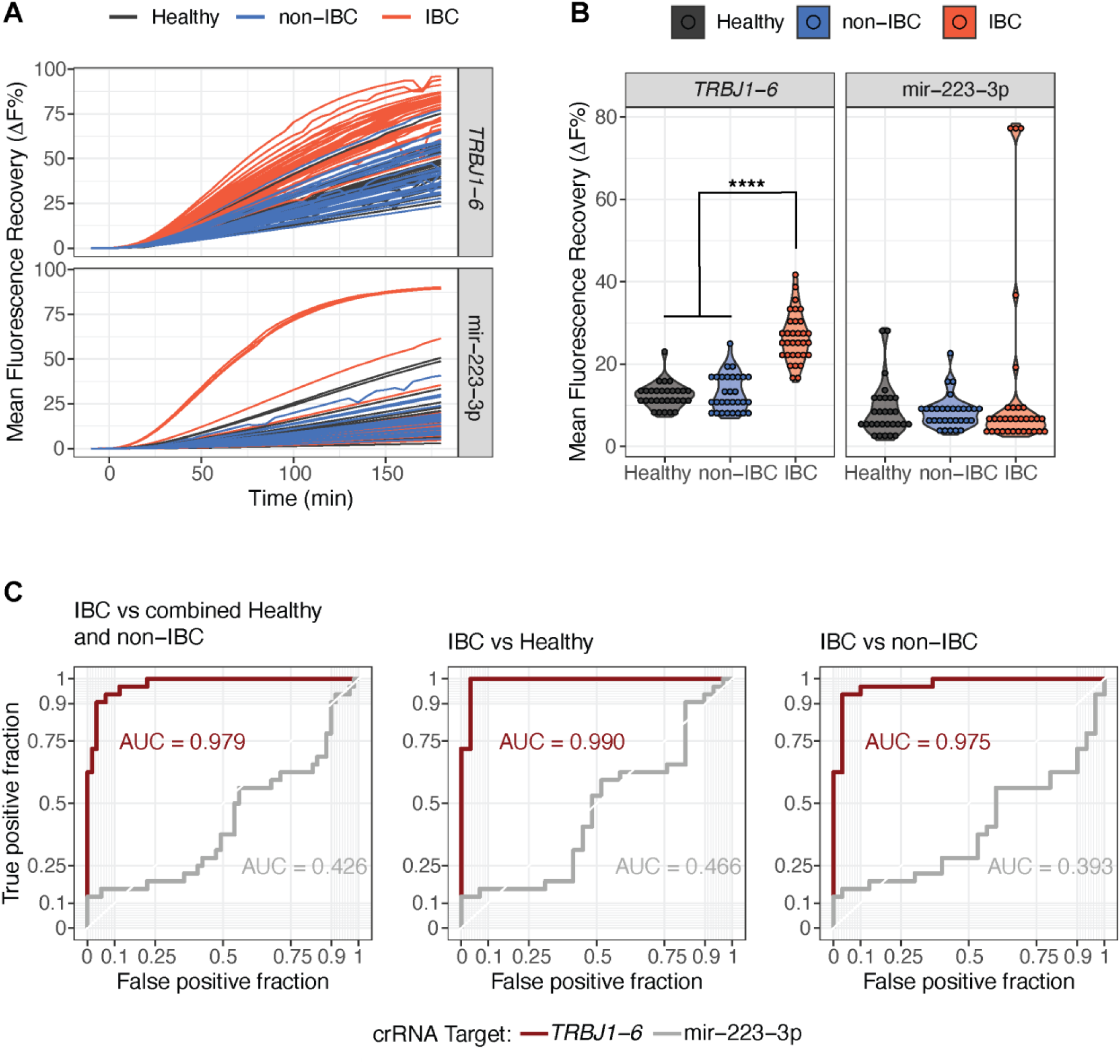
*TRBJ1-6* biomarker RT-PCR/Cas12a assay for IBC patient PBMCs compared to healthy donor and non-IBC patient PBMCs. RT-PCR/Cas12a assay detection of the *TRBJ1-6* fragment and mir-223-3p was done for two technical replicates of PBMC RNAs from 32 IBC patients, 30 non-IBC patients, and 29 healthy donors. (**A**) RT-PCR/Cas12a assay detection over 180-min time courses for each sample used to calculate the Mean Fluorescence Recovery (ΔF%) from two technical replicates. **(B)** Violin plots comparing detection of each target RNA at optimal time points of 65 min for *TRBJ1-6* and 110 min for mir-223-3p based on maximum Area Under the Curve (AUC) values in Figure S2A. The differences in *TRBJ1-6* levels in IBC patient PBMCs were highly significant (****) compared to those in healthy donor (p_adj_ = 1.15E-10) and non-IBC patients (p_adj_ = 4.79E-10). Differences in mir-223-3p levels in these samples were not statistically significant. Statistics were determined by a Kruskal-Wallis rank sum test with a post-hoc Dunn’s test with a Holm-Bonferroni correction. **(C)** Receiver Operating Curve (ROC) analysis of data in panels A and B, indicated a high diagnostic potential of the *TRBJ1-6* RNA in IBC patient PBMCs compared to a combined group of healthy donor and non-IBC patient PBMCs (*TRBJ1-6* AUC = 0.979, at the 65-min time point). mir-223-3p contrasts as a control for the assay, but not an IBC diagnostic biomarker (mir-223-3p AUC = 0.426 at the 110-min time point). The same trends were found in IBC patient PMBCs compared to either healthy donor PBMCs (*TRBJ1-6* AUC = 0.990, mir-223-3p AUC = 0.466, corresponding to 25-min time points for both targets) or non-IBC patient PBMCs (*TRBJ1-6* AUC = 0.975, mir-223-3p AUC = 0.393, at the 180-min time points for both targets). The time points chosen for displaying ROC curves were determined by time-course of AUC data in Figure S2, as the time point with the maximum AUC, although the AUC was relatively stable across the entire time course for both targets.

### Factors that potentially contribute to the generation of IBC *TRBJ1-6* RNA fragments in PBMCs

The findings above indicated that the overall abundance and stability of the *TRBJ1-6* RNA fragment in IBC PBMCs likely reflects the presence of a 2’,3’cP, in line with other sequencing studies that noted the prevalence of RNAs with PNK-removable 2’,3’cPs (mRNA and lncRNA fragments and 5’ tRNA halves) as putative biomarkers in human plasma and ER+ luminal breast cancer FFPE tissues (*16*, *18*, *19*, *21*, *23*). To identify factors that might contribute to the accumulation of this RNA fragment in PBMCs, we analyzed previously obtained TGIRT-seq datasets for IBC patient versus non-IBC patient and healthy donor PBMCs with reads mapped to the genome (Exons + Introns) and transcriptome (Exons only) reference sequences (Figure 5). Previous analysis of the TGIRT-seq datasets in this manner suggested pervasive enhanced transcription in highly activated IBC patient PBMCs that for many genes results in rate-limiting RNA splicing, leading to the accumulation of unspliced pre-mRNAs and impacting levels of mature mRNA (*7*).

**Figure 5.**
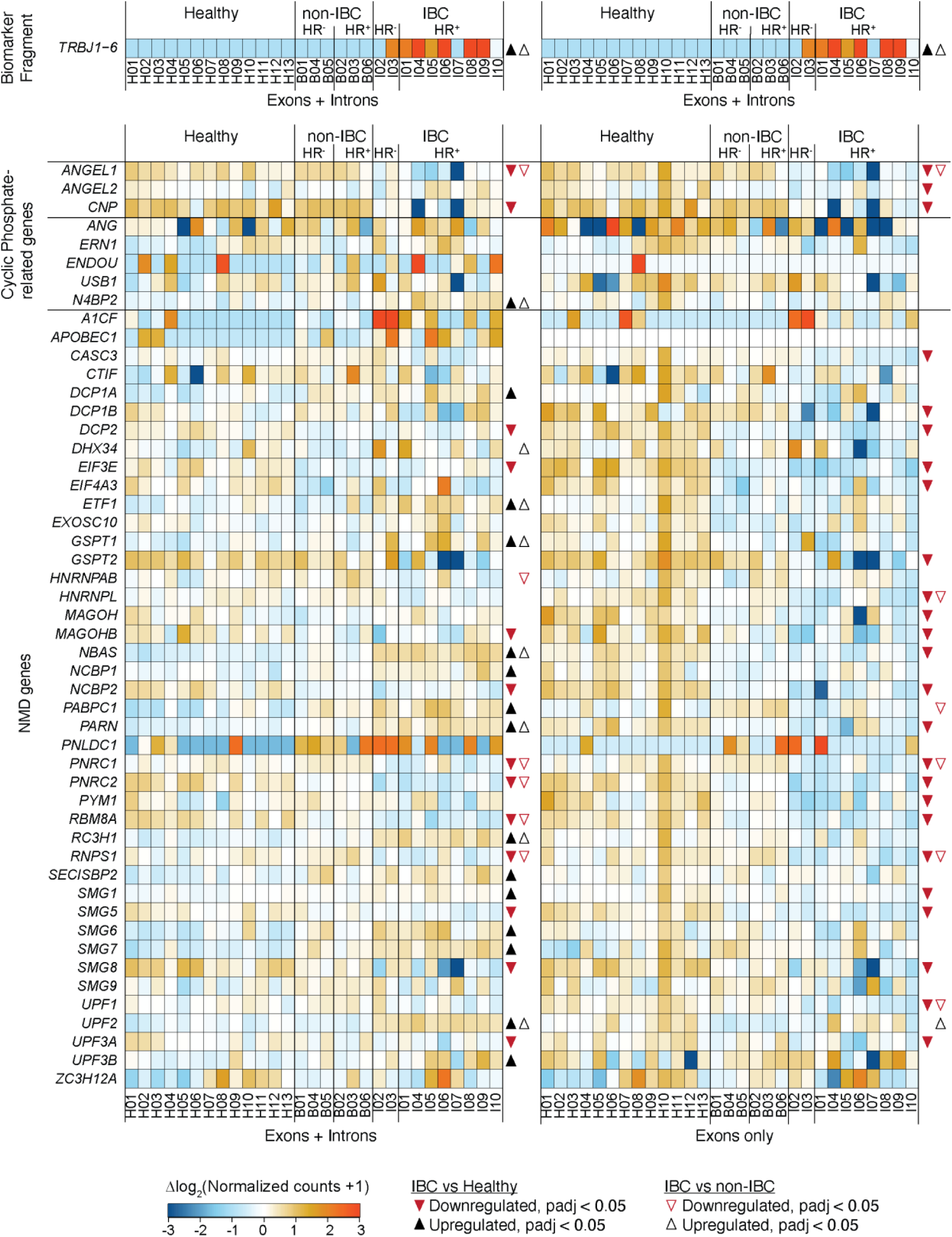
*TRBJ1-6* levels in PBMCs may reflect interplay of gene expression changes in cyclic phosphatase removal and NMD pathway genes. Heat maps showing TGIRT-seq quantification of healthy donor, non-IBC patient, and IBC patient PBMC RNAs from previous TGIRT-seq datasets (*7*) with reads mapped to genome (Exons + Introns, left) or transcriptome (Exons only, right) and read counts normalized using DESeq2. The *TRBJ1-6* RNA fragment expression is included only for paired-end read lengths ≤30 nt mapped to the genome reference sequence (Exons + Introns), since it includes intronic sequences (expression levels duplicated above each heat map for easier comparisons). The top two gene groups show cyclic phosphate related genes; 2’,3’-cyclic phosphatases (*ANGEL1*, *ANGEL2, CNP*) (*27–30*), and endo- and exonucleases that generate 2’,3’cP (*ANG*, *ERN1*, *ENDOU*, *USB1*, *N4BP2*) (*16*, *29*). The bottom group shows Nonsense-Mediated RNA Decay (NMD) genes from the Gene Ontology (GO) term “nuclear-transcribed mRNA catabolic process, nonsense-mediated decay” (GO ID GO:0000184), supplemented with recently described cell-type specific NMD factor *HNRNPL* (*31–33*). Individual tiles are color-coded as shown below the heat map for normalized counts transformed as the difference relative to the genewise mean taken across all PBMC samples for that gene. Lanes are labeled at the bottom according to the healthy donor and patient ID numbers from Wylie *et al.* (*7*). IBC and non-IBC patient samples are grouped as hormone receptor positive (HR+) or hormone receptor negative (HR-). Statistical comparisons between IBC and healthy (closed triangles) and IBC and non-IBC (open triangles) are notated by symbols to the right of the heat maps for those with p_adj_ <0.05, calculated by DESeq2 (symbols red if significantly downregulated in IBC and black if significantly upregulated in IBC compared to healthy donors (filled in triangles) or non-IBC patients (open triangles)).

Analysis of TGIRT-seq datasets with reads mapped to the transcriptome reference sequence (Exons only) indicated that PBMCs in most IBC patients had lower levels of mRNAs than healthy donor PBMCs for three genes that encode 2’,3’-cyclic phosphatases, *ANGEL1*, *ANGEL2*, and *CNP* (*27–30*) (Figure 5, right panel), which may lead to higher levels of 2’3’-cyclic phosphates on *TRBJ1-6* RNA fragments that contribute to their higher levels and stability in IBC patient PBMCs and plasma. Comparisons with reads mapped to the genome reference sequence suggested that the lower levels of mRNAs for *ANGEL1* and *CNP* mRNAs in IBC patient PBMCs was due to decreased transcription (lower levels in IBC patients for reads mapping to both Exon + Introns and Exons only) and rate-limiting RNA splicing for *ANGEL2* (lower levels in IBC patients for reads mapping to Exons only but not for reads mapping to Exons + Introns; Figure 5, compare left and right panels). By contrast, mRNA levels for four endonucleases and one exonuclease that generate 2’,3’cP modifications (*ANG*, *ERN1* (*IRE1*), *ENDOU* (*PP11*), *N4BP2*, and *USB1*) (*16*, *29*) were not consistently different in IBC patient versus non-IBC patient and healthy donor PBMCs for reads mapped to the transcriptome reference sequences (Figure 5).

Nonsense-mediated RNA decay (NMD) (*31–33*), which degrades mRNAs with premature termination codons (PTCs), is another pathway that could impact the abundance of *TRBJ1-6* RNA fragments in IBC patient PBMCs. Programmed DNA recombination at the *TCR*β locus is highly error prone, with two out of every three recombination events leading to frameshift mutations that could create PTCs, which trigger robust downregulation by NMD (*31*, *34*, *35*). Knockdown of key NMD factors, UPF1 (part of the core NMD machinery) and eIF4AIII (exon-junction complex protein that participates in NMD), both led to >3-fold upregulation of PTC containing TCRβ reporters compared to a control TCRβ reporter without a PTC (*34*). TCRβ mutations and frameshifts have been documented to lead to inhibited splicing and pre-mRNA accumulation due to the gene’s unique density of exonic-splicing enhancers (*36*), and NMD disruption in T cell development in mice, through T-cell specific *UPF2* deletion, led to the accumulation of nonproductively rearranged by-products of TCRβ transcripts (*37*). The NMD pathway plays an important role in regulating the expression of both cytokine and cytokine receptor mRNAs during inflammatory responses (*38*, *39*) and downregulated NMD as a result of *UPF1* mutations has been linked to inflammatory myofibroblastic tumors (*40*).

Analysis of the previous TGIRT-seq datasets showed that many genes involved in the NMD pathway have significantly lower mRNA expression levels in IBC versus healthy donor PBMCs and in some cases also versus non-IBC patient PBMCs for reads mapped to the transcriptome reference sequence (Exons only). These include key NMD factors: *UPF1* and *UPF3A* (components of the core NMD machinery); *EIF4A3*, *CASC3*, *RBM8A*, *MAGOH*, *RNPS1*, and *PYM1* (Exon-Junction complex proteins that function in NMD); *SMG1* (kinase that phosphorylates the UPF1 protein) and *SMG5* (factor that recruits phosphatases to promote dephosphorylation of the UPF1 protein) (Figure 5) (*31*). These findings suggest a model that down regulation of NMD repression in IBC prevents the normally robust degradation of nonproductively rearranged TCRβ RNAs, potentially contributing to the accumulation of *TRBJ1-6* and other *TRBJ1* fragments. Further experiments will be needed to assess how differences in protein expression levels of cyclic phosphatases or NMD proteins impacts the prevalence of the *TRBJ1-6* RNA biomarker.

## Discussion

Our findings support the previous TGIRT-seq identification of the *TRBJ1-6* pre-mRNA fragment as a blood-based biomarker for IBC by using a precision high-throughput RT-PCR/Cas12a detection method, enhanced to detect 2’,3’cP RNAs by the use of PNK treatment. Comparing PBMC RNAs from 32 IBC patients, 30 non-IBC patients, and 29 healthy donors, we found *TRBJ1-6* was significantly enriched compared to healthy donor (p_adj_ = 1.15E-10) and non-IBC patients (p_adj_ = 4.79E-10) with ROC analysis for IBC patients compared to combined healthy donors and non-IBC patients having an AUC = 0.979, indicating that this blood-based biomarker is highly promising for differentiating IBC from healthy donors and non-IBC patients (Figure 4). Our previous study (*7*) described how TGIRT-seq could detect numerous potential RNA biomarkers in IBC patient tumors, PBMCs, and plasma, and this study supports the previous identification of *TRBJ1-6* as a potential IBC biomarker by a high-throughput assay that could be used for a more rapid and less expensive initial diagnostic test than highly discriminatory TGIRT sequencing.

A PBMC origin for the plasma-detected biomarker is not surprising as cell-free blood-based RNAs (cfRNA) are predominantly derived from blood cells and platelets (*41*, *42*), and PBMCs have been shown previously to be a source of cancer-specific biomarkers (*43*). Although the biomarker was initially discovered in plasma, PBMCs may prove to be a more reliable source of the biomarker, as the yield of RNA isolated from a PBMC preparation is much higher than from plasma, providing enough material for more technical replicates and controls. Going forward, additional comprehensive TGIRT-seq of healthy donor and non-IBC and IBC patient PBMCs will be used to further assess gene expression differences that may contribute to *TRBJ1-6* expression and stability in IBC patient PBMCs, and additional high-throughput assays will be done to confirm other potential biomarkers identified by TGIRT-seq (*7*). This would further enhance the already high confidence levels of the current assay and provide additional insights into distinguishing complex differences between breast cancer subtypes that could impact personalized medicine for individual patients.

There are several possible explanations for the higher levels of the *TRBJ1-6* RNA biomarker in IBC patient PBMCs. The simplest is that the biomarker is increased in expression in the IBC patient PBMCs, most likely arising from T-cells. Another possible explanation is that the biomarker is expressed in neutrophils or eosinophils, whose activation in IBC was suggested by higher expression levels of other neutrophil and eosinophil markers identified by CIBERSORTx profiling of TGIRT-seq PBMC data (*7*). Although these granulocyte cell types are usually depleted from PBMC preparations due to their higher density, activation can change their density to allow their isolation with the lower density PBMC fraction (*44–46*). Productive protein expression of αβTCR has been described in a subpopulation of neutrophils, while a subpopulation of eosinophils was only found to express γδTCR proteins, so a neutrophil origin seems more likely than from eosinophils (*47–49*). A final possibility is that this biomarker is present in circulating tumor cells (CTCs) (*50*, *51*). It has already been well-established that cancer cells can express immunoglobulin receptors (*52*) and TCR gene expression has been found in diverse immortalized cell lines unrelated to the immune system (Human Protein Atlas (proteinatlas.org) (*53*, *54*). The same *TRBJ1-6* RNA fragment was detected by TGIRT-seq in IBC patient FFPE tumor samples and at lower levels in frozen non-IBC patient tumor and contralateral normal breast tissues tumor samples (*7*). This tumor expression suggests the *TRBJ1-6* fragment could also come from expression in CTCs, though detection in tumor samples could also arise from tumor-infiltrating lymphocytes. Performing detection assays in cell populations separated by flow cytometry could lead to a more robust understanding of the mechanism of this biomarker’s generation and IBC’s influence on the immune repertoire.

As RNA sequencing methods are refined for discovering a diversity of RNA biomarkers of diseases, care must be taken to fully characterize RNAs by size and specific sequence rather than using standard RNA-seq bioinformatic pipelines that combine gene count reads regardless of location in the RNA or length. The *TRBJ1-6* biomarker was discovered in plasma without needing to consider its length, whereas in PBMCs, only the short fragment was differentially expressed but not the full-length pre-mRNA. Adding length considerations to bioinformatic analysis increases the flexibility of detection of the biomarker from PBMCs and indicated a mechanism for the origination of the mRNA fragment biomarkers, as the same fragment found in plasma could be traced to PBMCs. It will be of interest to see if other cyclic phosphate RNA fragments serve as stable biomarkers for IBC and other diseases. Such RNA fragments may be more prevalent in cases where PBMCs have downregulated cyclic phosphatases and NMD related genes as found here for IBC. mRNA fragments with 2’,3’-cyclic phosphates may be particularly prevalent in diseases such as leukemia, autoimmunity, and inflammatory cancers for which specific classes of PBMCs are activated.

Although our RT-PCR/Cas12a *TRBJ1-6* detection assay performed well in distinguishing IBC patients from non-IBC patients and healthy donors, we note several limitations. First, there was some overlap between the lowest levels of *TRBJ1-6* RNA in IBC patient PBMCs and its highest level in non-IBC patient and healthy donor PBMCs. Notably, the non-IBC patient with the highest levels of the *TRBJ1-6* RNA detected by the RT-PCR/Cas12a assay was initially presented to the clinic with an IBC diagnosis, but ultimately through careful assessment was considered non-IBC and responded well to standard breast cancer treatments. At present, patients with *TRBJ1-6* RNAs in overlapping ranges could be singled out for further diagnostic tests including comprehensive TGIRT-seq to assess other differentially expressed RNAs that were found in IBC patient tumors, PBMCs, and plasma samples but have not yet been confirmed as biomarkers (*7*). Second, although *TRBJ1-6* levels distinguish IBC from non-IBC patient and healthy donor PBMCs, we did not yet assess how IBC patient PBMCs might differ from the most frequent IBC misdiagnosis, mastitis, a common breast inflammation that is often caused by bacterial infection and impedes rapid diagnosis of IBC (*6*, *55*). After further clinical validation, patients with undiagnosed inflammatory breast symptoms might benefit from incorporation of high-throughput assays of the *TRBJ1-6* RNA fragment or other TGIRT-seq identified differentially expressed RNAs into the current clinical algorithms (*7*). Finally, although robust, our high-throughput assay has many successive enzymatic steps, and there may be other ways to detect the *TRBJ1-6* biomarker with fewer manipulation steps. Overall, our TGIRT-seq analysis provides a roadmap for further understanding and diagnosing IBC, including unearthing and validating additional RNA biomarkers that may not be detectable by other sequencing methods. TGIRT-seq may provide similar advantages for understanding and diagnosing other diseases.

## Materials and Methods

### Patients and sample preparation

Patient samples were obtained and processed at MD Anderson Cancer Center following institutional protocols, as previously described (*7*). The MDACC Institutional Review Board approved this study and patients and healthy donors each gave written informed consent. Fresh blood was collected and PBMCs were isolated using Ficoll-Paque PLUS (GE Healthcare) density gradient centrifugation, following the manufacturer’s guidelines. Total RNA was extracted from PBMC samples at MD Anderson using the miRVana miRNA isolation kit (ThermoFisher) then shipped to UT Austin on dry ice and stored at -80°C.

### DNA and RNA oligonucleotides

DNA and RNA oligonucleotides were purchased from Integrated DNA Technologies (IDT) and are shown in Table S1. DNA oligonucleotides include universal reverse transcription primer, universal reverse PCR primer, and specific forward PCR primers (*8*, *56*). The dsDNA probe was composed of a 5’ 6-FAM (Fluorescein) labeled fluorophore reporter strand and 3′-IowaBlack labeled quencher reporter strand (*13*). RNA oligonucleotides include CRISPR RNAs (crRNA).

### PBMC RT-PCR/Cas12a assay

Phosphatase treatment (either T4 Polynucleotide Kinase (PNK) or Calf Intestine Phosphatase (CIP)) and the combined poly(A) addition and reverse transcription steps were performed in succession in the same PCR tube. In the first step, 20 ng of PBMC RNA for each patient or healthy donor sample were incubated with PNK to remove 3’-phosphates under the following conditions: 10 U T4 Polynucleotide Kinase (New England Biolabs), 20 U RNase Inhibitor, Murine (New England Biolabs) in 1X T4 Polynucleotide Kinase Reaction Buffer (10-μL final reaction volume) for 30 min at 37°C followed by 20 min at 65°C. In a parallel test reaction to assess presence of a 2’,3’-cyclic phosphate versus 3’-phosphate, 20 ng of PBMC RNA was incubated with CIP instead of PNK: 5 U Quick CIP (New England Biolabs), 20 U RNase Inhibitor, Murine (New England Biolabs), in 1X rCutSmart Buffer (10-μL final reaction volume) for 15 min at 37°C and 2 min at 80°C. Following either phosphatase treatment, the addition of poly(A)-tails and reverse transcription were performed simultaneously by adding an additional 10 μL of a reaction mixture (2.5 U *E. coli* Poly(A) Polymerase (New England Biolabs), 100 U Maxima H Minus Reverse Transcriptase (ThermoFisher), 20 U RNase Inhibitor, Murine (New England Biolabs), 0.5 mM dNTP, 1 mM ATP, 100 pmol universal reverse transcriptase (RT) primer, in 1X Maxima RT Buffer) and incubated for 1 min at 37°C, 30 min at 50°C, and 5 min at 85°C. Control reactions omitting each individual enzyme were performed under the same conditions.

Following the above treatment, PCRs were performed in a standard thermal cycler using TaqMan Fast Advanced Master Mix for qPCR (ThermoFisher) with the optional uracil-N-glycosylase (UNG) step to prevent PCR carry-over contamination (2 μL cDNA, 800 nM specific forward primer, 800 nM universal reverse primer (*8*), 80 ng ET SSB (Extreme Thermostable Single-Stranded DNA Binding Protein) (New England Biolabs) in 20 μL final reaction volume; incubated for 2 min at 50°C, 2 min at 95°C, 25 to 35 cycles: 3 s at 95°C, 30 s at 60°C). The number of PCR cycles was optimized for each target to prevent too early signal saturation which results in photobleaching and signal decrease over time (35 cycles for *TRBJ1-6* and 25 cycles for mir-223-3p).

The final CRISPR-detection step was a modification of that described in Smith *et al*. (*13*). Reporter reaction buffer consisted of a probe fluorophore reporter strand (0.3 μM; Table S1) and a quencher reporter strand (0.3 μM; Table S1) in 1X CRISPR buffer (New England Biolabs Buffer r2.1 or a similar buffer (10 mM Tris-HCl (pH 7.9), 50 mM NaCl, 10 mM MgCl_2_, 100 μg/mL bovine serum albumin) in a final volume of 96 μL per sample detection reaction (Table S1). The Reporter reaction buffer was incubated protected from light for 30 min at room temperature and then aliquoted (96 μL) into a 96-well black, clear bottom plate (Falcon). PCR amplicons were added to the plate with a 4-μL input. Target specific CRISPR RNAs (0.72 μM) were incubated with EnGen Lba Cas12a (Cpf1) enzyme (0.6 μM, New England Biolabs or ThermoFisher) in 1X CRISPR buffer for 30 min at 37°C. The plate was inserted into a Clariostar Plus plate reader (BMG Labtech) for measurements with Enhanced Dynamic Range (EDR) settings at 37°C, with bottom optic measurements (filter settings, excitation: 482-16, dichroic: LP 504, emission: 530-40; shaking before each cycle, double orbital 400 rpm 10 s before each cycle). Background readings were taken first (11 cycles, 60 s each, 50 flashes per well). After background readings were completed, 5 μL pre-incubated Cas12a/crRNA complex was added to each well and fluorescence intensity measured every 5 min for the next 180 min (36 cycles, 300 s each, 50 flashes per well). Fluorescence recovery (ΔF%) was calculated using the equation from Smith *et al.* (*14*): (F_obs_ − F_q)_/(F_max_ − F_q_) × 100%, where F_obs_ is the observed fluorescence at each time point, F_q_ is the quenched fluorescence of the assembled probe (determined as the mean of the background readings prior to the addition and activation of the crRNA-Cas12a complex), and F_max_ is the maximum fluorescence of the probe fluorophore reporter strand (added without the quencher reporter strand), included as a control well on each plate. Each sample was assayed in duplicate technical replicates to calculate the Mean Fluorescence Recovery (ΔF%).

### Data analysis

TGIRT-seq datasets were previously generated in Wylie *et al.* (*7*) and deposited in the National Center for Biotechnology Information Sequence Read Archive (accession number: PRJNA954747). Mapped datasets for PBMCs were filtered using SAMtools v1.20 with customized settings for filtering the reads to reads corresponding to RNAs that were 30 nt or shorter (samtools view -h -f 67 -e ‘length(seq)<=30 && (length(seq)==tlen || length(seq)==-tlen)’). DESeq2 (*57*) was used for normalization and statistical analysis for comparisons of filtered protein-coding reads in TGIRT-seq datasets, as described in Wylie *et al.* (*7*). Integrated Genomics Viewer (IGV) version 2.17.4 was used for visualizing read coverage of TGIRT-seq datasets (*58*). Receiver operating characteristic (ROC) curves and area under the curve (AUC) analyses were generated using the R package “plotROC” (*59*).

### Statistical Analysis

All visualizations and data analysis were performed using R (4.3.3) and the “dplyr” R package (*60*, *61*). Statistics for Figure 4B violin plots, corresponding to large-scale RT-PCR/Cas12a detection assays, were determined by Kruskal-Wallis rank sum test (*60*) with pairwise comparisons determined by post-hoc Dunn’s test with a Holm-Bonferroni correction for multiple comparisons (*62*). Statistical comparisons were determined using the R packages “stats” and “FSA” (*60*, *62*).

## Data Availability

All data produced in the present study are available upon reasonable request to the authors.

## Acknowledgments

This work represents translational science from samples prospectively collected and annotated after patient specific consent on an IRB approved registry from patients recruited from the dedicated IBC clinic at MDACC. A large group of people, both patient-facing and behind the scenes, are members of the author-listed MDACC Inflammatory Breast Cancer Team that made this translational science possible. They include Rachel Layman, Bora Lim, Sadia Saleem, Vicente Valero, Michael C. Stauder, Anthony Lucci, Susie X. Sun, Gary J. Whitman, Miral Patel, Huong Le-Petross, Yang Lu, Angela Marx, Angela Alexander, Chasity Yajima, Megumi Kai, Lily Villarreal, Heather Lopez. We thank the patients and healthy donors for contributing samples and the staff of the Morgan Inflammatory Breast Cancer Research Program and Clinic for collecting those samples. We also thank Philomena Alapatt (University of Texas at Austin) for comments on the manuscript. The sequencing of the TGIRT-seq libraries in Wylie *et al.* (*7*) that provided additional insights in the present manuscript was done by the Genomic Sequencing and Analysis Facility at UT Austin, Center for Biomedical Research Support (CBRS). The CBRS at the University of Texas at Austin also provided high-performance computing resources.

## Funding

National Institutes of Health grant R35 GM136216 (AML)

National Institutes of Health /National Cancer Institute grant 1R01CA284102 (WAW)

National Institutes of Health /National Cancer Institute grant R01 CA264529-01 (WAW)

Susan G. Komen grant OG250001 (WAW)

Breast Cancer Research Foundation CONS-23-010 (WAW)

The State of Texas Grant for Rare and Aggressive Breast Cancer (WAW)

National Institutes of Health /National Cancer Institute grant 4P30CA016672-48 (WAW)

National Institutes of Health grant 1RO1CA284102 (SK)

National Institutes of Health grant 1RO1CA264529-01 (SK)

National Institutes of Health grant 1RO1CA258523 (SK and NTU)

Breast Cancer Research Foundation grant BCRF-22-164 (NTU)

## Author contributions

Conceptualization: EAF-K, CWS, NTU, AML

Methodology: EAF-K, SD, CWS

Software: EAF-K, JY

Validation: EAF-K, SD

Formal analysis: EAF-K, JY

Investigation: EAF-K, SD

Resource: WAW, SK, NTU, AML, The MDACC Inflammatory Breast Cancer Team

Data curation: EAF-K, JY

Visualization: EAF-K, JY

Supervision: WAW, SK, NTU, AML

Writing—original draft: EAF-K, AML

Writing—review & editing: EAF-K, SD, JY, CWS, XW, WAW, NTU, SK, AML

Project administration: WAW, SK, NTU, AML

Funding acquisition: WAW, SK, NTU, AML

## Competing interests

AML is an inventor on patents owned by the University of Texas at Austin for TGIRT enzymes and other stabilized reverse transcriptase fusion proteins and methods for non-retroviral reverse transcriptase template switching. EAF-K, JY, XW, NTU, and AML are listed inventors on a patent application filed jointly by UT Austin and MD Anderson entitled “Methods and Compositions for Diagnosing, Treating and/or Preventing Inflammatory Breast Cancer”, including the RNA biomarker confirmed in this study. CWS is listed as an inventor on a patent filed by the University of Albany, State University of New York for Cas12a trans-cleavage of dsDNA reporters and methods for optimizing the reporting rate. All other authors declare that they have no competing interests.

## Data and materials availability

The TGIRT-seq datasets in this manuscript were generated previously (*7*) and deposited in the National Center for Biotechnology Information Sequence Read Archive (accession number: PRJNA954747). Upon reasonable request to Dr. Krishnamurthy, deidentified patient data will be made available after Institutional Review Board approval by MD Anderson. All data produced in the present study are available upon reasonable request to the authors.

## Notes

### Author Declarations

The MDACC Institutional Review Board approved this study and patients and healthy donors each gave written informed consent.

