## Supplementary Figures and Tables for "High-throughput assay confirmation of a T-cell receptor pre-mRNA fragment as a blood-based inflammatory breast cancer biomarker"

Elizabeth A. Ferrick-Kiddie *et al.*

**This PDF file includes:**

Figures S1 and S2
Tables S1 and S2

**Figure S1**

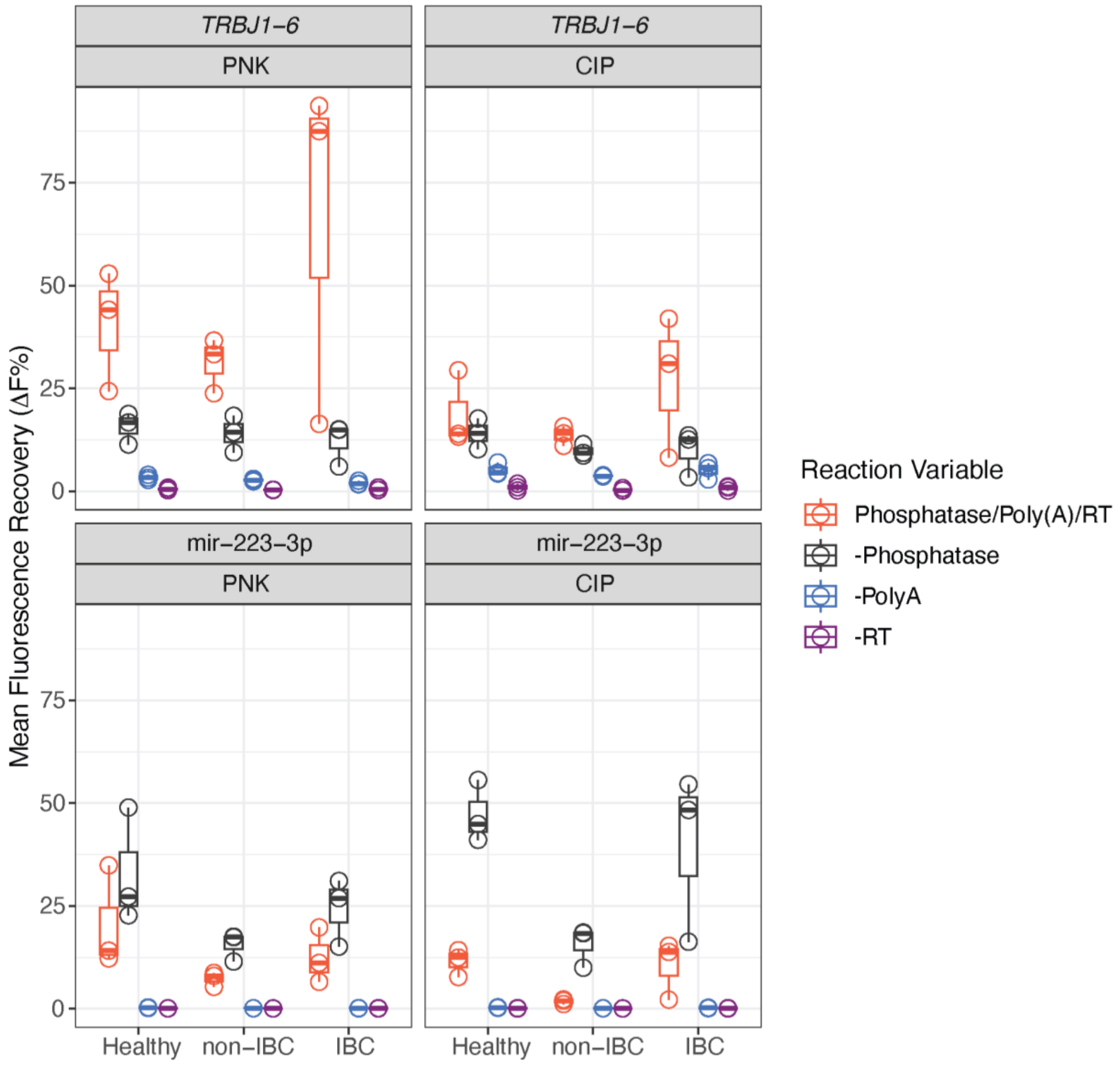


**Figure S1.** ***TRBJ1-6* biomarker detection with low background in the RT-PCR/Cas12a diagnostic assay.** Optimal detection of *TRBJ1-6* in PBMC samples is dependent upon PNK treatment instead of CIP treatment. Control reactions omit a phosphatase (-Phosphatase, either PNK or CIP), Poly(A) polymerase (-Poly(A)), or reverse transcriptase (-RT) in the RT-PCR/Cas12a detection assay. Each PBMC sample group was composed of two technical replicates for three patient sample PBMCs to calculate the Mean Fluorescence Recovery (ΔF%). The 180-min timepoint was chosen as representative for the comparison.

**Figure S2**

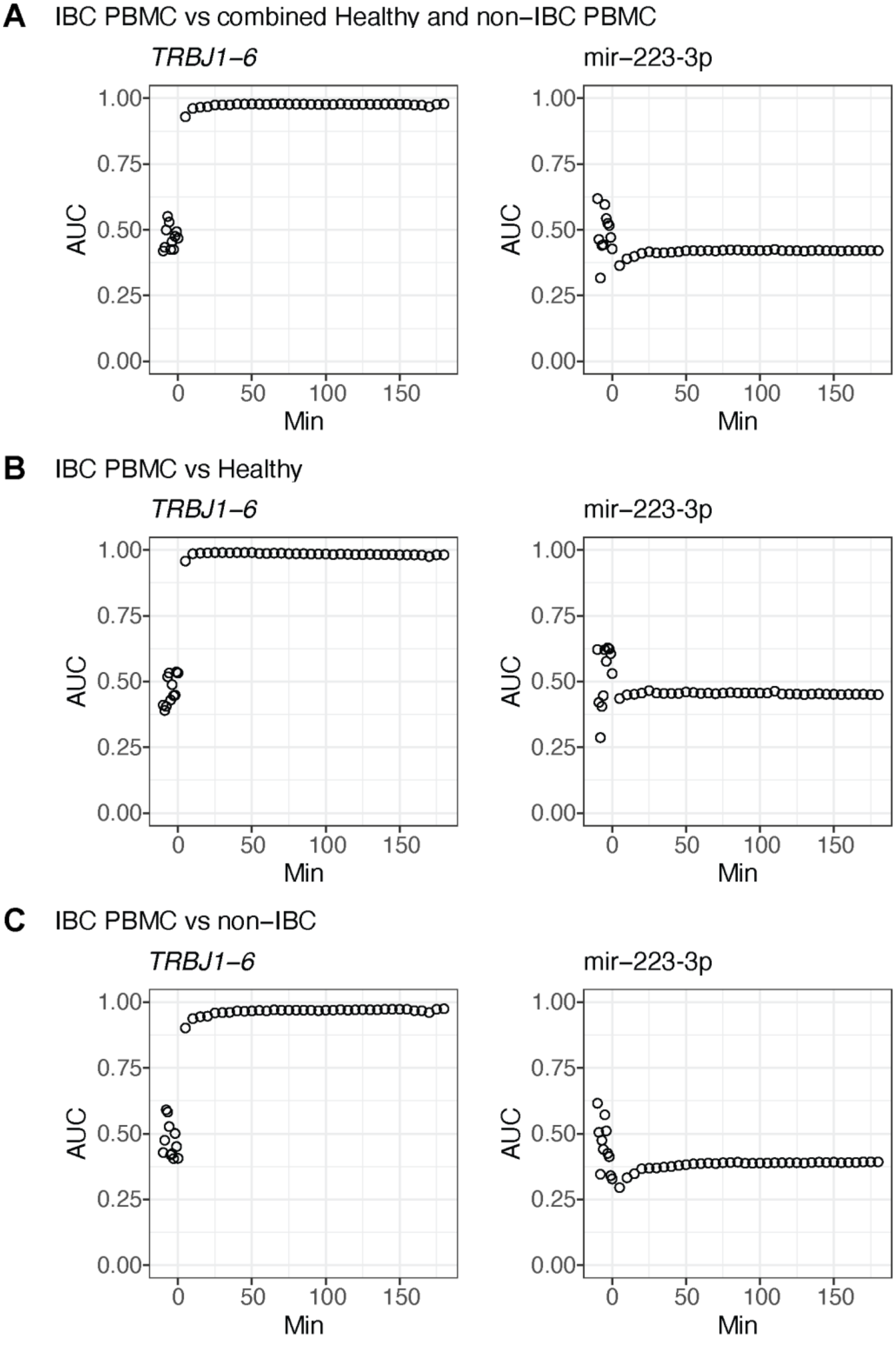

A
IBC PBMC vs combined Healthy and non−IBC PBMC
TRBJ1−6
mir−223-3p
AUC
1.00
0.75
0.50
0.25
0.00
0
50
100
150
Min
B
IBC PBMC vs Healthy
TRBJ1−6
mir−223-3p
C
IBC PBMC vs non−IBC
TRBJ1−6
mir−223-3p

**Figure S2. AUC measurements are stable across a 180-min time period in the RT-PCR/Cas12a diagnostic assay.** Area Under the Curve (AUC) measurements at each of the timepoints during the 180 min RT-PCR/Cas12a diagnostic assay are graphed, including the baseline measurements prior to Cas12a-crRNA addition, which show that the assay only becomes specific after Cas12a-crRNA addition. Although we chose the timepoint with the highest AUC for Figure 4 graphs, the AUC was stable across the entire time course for both targets. AUC time courses are shown for *TRBJ1-6* and mir-223-3p for the following group comparisons: (A) IBC patient PBMCs versus combined healthy donor and non-IBC patient PBMCs; (B) IBC patient PBMCs versus healthy donor PBMCs; and (C) IBC patient PBMCs versus non-IBC patient PBMCs.

**Table S1: DNA and RNA Oligonucleotides**

| Name | Sequence |
|---|---|
| Universal RT primer* | CAGTGCAGGGTCCGAGGTTTTTTTTTTTTTTTTTTTVN |
| Universal reverse PCR primer* | CAGTGCAGGGTCCGAGGT |
| TRBJ1-6 forward PCR primer | TTGACCAGGCTCACTGTG |
| hsa-mir-223-3p forward PCR primer* | CACGGTGTCAGTTTGTCAA |
| Probe fluorophore reporter strand† | /56-FAM/AGAACCGAAGTGTAGCTTATCA |
| Probe quencher reporter strand† | TGATAAGCTACACTTCGGTTCT/3IABkFQ/ |
| TRBJ1-6 crRNA | rUrArArUrUrUrCrUrArCrUrArArGrUrGrUrArGrArUrCrArUrArC rCrUrGrUrCrArCrArGrUrGrArGrCrCrU |
| Hsa-mir-223-3p crRNA | rUrArArUrUrUrCrUrArCrUrArArGrUrGrUrArGrArUrUrGrGrGr GrUrArUrUrUrGrArCrArArArCrUrGrArC |

*Based on Kang *et al.,* 2012 and Niu *et al*., 2015

†Based on Smith *et al*., 2021

### Table S2: Patient and Healthy Donor Information

| Donor ID | Sample Type | Age | Race | Ethnicity | Smoking | ER+/- | PR+/- | HER2+/- | Histology | Nuclear Grade | TNM | Stage | Subtype |
|---|---|---|---|---|---|---|---|---|---|---|---|---|---|
| H01 | Healthy PBMC | 71-75 | Black or African American | Not Hispanic or Latino | No, never | NA | NA | NA | NA | NA | NA | NA | NA |
| H02 | Healthy PBMC | 51-55 | White or Caucasian | Not Hispanic or Latino | No, never | NA | NA | NA | NA | NA | NA | NA | NA |
| H03 | Healthy PBMC | 61-65 | Other | Hispanic or Latino | No, never | NA | NA | NA | NA | NA | NA | NA | NA |
| H04 | Healthy PBMC | 61-65 | White or Caucasian | Not Hispanic or Latino | No, never | NA | NA | NA | NA | NA | NA | NA | NA |
| H05 | Healthy PBMC | 31-35 | Asian | Not Hispanic or Latino | No, never | NA | NA | NA | NA | NA | NA | NA | NA |
| H06 | Healthy PBMC | 71-75 | White or Caucasian | Not Hispanic or Latino | Former | NA | NA | NA | NA | NA | NA | NA | NA |
| H07 | Healthy PBMC | 66-70 | White or Caucasian | Not Hispanic or Latino | No, never | NA | NA | NA | NA | NA | NA | NA | NA |
| H08 | Healthy PBMC | 36-40 | White or Caucasian | Not Hispanic or Latino | No, never | NA | NA | NA | NA | NA | NA | NA | NA |
| H09 | Healthy PBMC | 46-50 | White or Caucasian | Hispanic or Latino | No, never | NA | NA | NA | NA | NA | NA | NA | NA |
| H10 | Healthy PBMC | 51-55 | White or Caucasian | Not Hispanic or Latino | No, never | NA | NA | NA | NA | NA | NA | NA | NA |
| H11 | Healthy PBMC | 66-70 | White or Caucasian | Not Hispanic or Latino | No, never | NA | NA | NA | NA | NA | NA | NA | NA |
| H12 | Healthy PBMC | 71-75 | White or Caucasian | Not Hispanic or Latino | No, never | NA | NA | NA | NA | NA | NA | NA | NA |
| H13 | Healthy PBMC | 61-65 | White or Caucasian | Not Hispanic or Latino | Yes, currently smoke | NA | NA | NA | NA | NA | NA | NA | NA |
| H14 | Healthy PBMC | 66-70 | White or Caucasian | Not Hispanic or Latino | Former | NA | NA | NA | NA | NA | NA | NA | NA |
| H15 | Healthy PBMC | 61-65 | White or Caucasian | Not Hispanic or Latino | No, never | NA | NA | NA | NA | NA | NA | NA | NA |
| H16 | Healthy PBMC | 66-70 | White or Caucasian | Not Hispanic or Latino | Former | NA | NA | NA | NA | NA | NA | NA | NA |
| H17 | Healthy PBMC | 66-70 | White or Caucasian | Hispanic or Latino | No, never | NA | NA | NA | NA | NA | NA | NA | NA |
| H18 | Healthy PBMC | 66-70 | White or Caucasian | Not Hispanic or Latino | No, never | NA | NA | NA | NA | NA | NA | NA | NA |
| H19 | Healthy PBMC | 71-75 | White or Caucasian | Not Hispanic or Latino | No, never | NA | NA | NA | NA | NA | NA | NA | NA |
| H20 | Healthy PBMC | 61-65 | White or Caucasian | Not Hispanic or Latino | No, never | NA | NA | NA | NA | NA | NA | NA | NA |

| | | | | | | | | | | | | | |
|---|---|---|---|---|---|---|---|---|---|---|---|---|---|
| H21 | Healthy PBMC | 66-70 | White or Caucasian | Not Hispanic or Latino | No, never | NA | NA | NA | NA | NA | NA | NA | NA |
| H22 | Healthy PBMC | 66-70 | Black or African American | Not Hispanic or Latino | No, never | NA | NA | NA | NA | NA | NA | NA | NA |
| H23 | Healthy PBMC | 80-85 | White or Caucasian | Not Hispanic or Latino | No, never | NA | NA | NA | NA | NA | NA | NA | NA |
| H24 | Healthy PBMC | 46-50 | White or Caucasian | Not Hispanic or Latino | Former | NA | NA | NA | NA | NA | NA | NA | NA |
| H25 | Healthy PBMC | 41-45 | White or Caucasian | Not Hispanic or Latino | No, never | NA | NA | NA | NA | NA | NA | NA | NA |
| H26 | Healthy PBMC | 66-70 | White or Caucasian | Not Hispanic or Latino | No, never | NA | NA | NA | NA | NA | NA | NA | NA |
| H27 | Healthy PBMC | 56-60 | White or Caucasian | Not Hispanic or Latino | Former | NA | NA | NA | NA | NA | NA | NA | NA |
| H28 | Healthy PBMC | 51-55 | White or Caucasian | Hispanic or Latino | No, never | NA | NA | NA | NA | NA | NA | NA | NA |
| H29 | Healthy PBMC | 56-60 | White or Caucasian | Not Hispanic or Latino | No, never | NA | NA | NA | NA | NA | NA | NA | NA |
| H30 | Healthy PBMC | 46-50 | White or Caucasian | Hispanic or Latino | No, never | NA | NA | NA | NA | NA | NA | NA | NA |
| H31 | Healthy PBMC | 61-65 | White or Caucasian | Not Hispanic or Latino | Former | NA | NA | NA | NA | NA | NA | NA | NA |
| I01 | IBC PBMC | 46-50 | White or Caucasian | Hispanic or Latino | Former | ++ | ++ | - | Ductal | Unknown | T4dN0M0 | IIIB | Luminal A |
| I02 | IBC PBMC | 51-55 | White or Caucasian | Not Hispanic or Latino | No, never | + | - | + | Ductal | 3 | T4dN3aM0 | IIIC | Luminal B |
| I03 | IBC PBMC | 41-45 | Other | Hispanic or Latino | No, never | ++ | ++ | - | Ductal | 2 | T4dN1M0 | IIIB | Luminal A |
| I04 | IBC PBMC | 51-55 | White or Caucasian | Not Hispanic or Latino | No, never | ++ | - | - | Ductal | 3 | T4dN3aM0 | IIIC | Luminal A |
| I05 | IBC PBMC | 41-45 | White or Caucasian | Not Hispanic or Latino | No, never | - | - | - | Ductal | 3 | T4dN3cM0 | IIIC | Triple negative |
| I06 | IBC PBMC | 51-55 | White or Caucasian | Not Hispanic or Latino | No, never | ++ | + | - | Ductal | 3 | T4dN3M0 | IIIC | Luminal A |
| I07 | IBC PBMC | 66-70 | White or Caucasian | Not Hispanic or Latino | No, never | ++ | - | - | Ductal | 2 | T4dN0M0 | IIIB | Luminal A |
| I08 | IBC PBMC | 41-45 | White or Caucasian | Not Hispanic or Latino | Former | ++ | ++ | - | Ductal | 3 | T4dN0M0 | IIIB | Luminal A |
| I09 | IBC PBMC | 71-75 | White or Caucasian | Not Hispanic or Latino | No, never | - | - | - | Ductal | 2 | T4dN1M0 | IIIB | Triple negative |
| I10 | IBC PBMC | 41-45 | White or Caucasian | Not Hispanic or Latino | Former | ++ | ++ | - | Ductal | 2 | T4dN3aM0 | IIIB | Luminal A |
| I11 | IBC PBMC | 41-45 | White or Caucasian | Not Hispanic or Latino | Former | - | - | - | Ductal | 3 | T4dN3M0 | IIIC | Triple negative |

| | | | | | | | | | | | | | |
|---|---|---|---|---|---|---|---|---|---|---|---|---|---|
| I12 * | IBC PBMC | 46-50 | White or Caucasian | Not Hispanic or Latino | No, never | - | - | - | Ductal | 3 | T4dN3aM0 | IIIC | Triple negative |
| I13 | IBC PBMC | 51-55 | White or Caucasian | Hispanic or Latino | No, never | - | - | - | Ductal | 3 | T4dN3bM0 | IIIC | Triple negative |
| I14 | IBC PBMC | 46-50 | White or Caucasian | Not Hispanic or Latino | No, never | ++ | + | - | Ductal | 3 | T4dN1M0 | IIIB | Luminal A |
| I15 | IBC PBMC | 51-55 | Black or African American | Not Hispanic or Latino | No, never | ++ | ++ | - | Ductal | 2 | T4dN3aM0 | IIIB | Luminal A |
| I16 | IBC PBMC | 61-65 | White or Caucasian | Not Hispanic or Latino | Former | - | - | - | Ductal | 3 | T4dN1M0 | IIIC | Triple negative |
| I17 | IBC PBMC | 71-75 | White or Caucasian | Not Hispanic or Latino | Former | - | - | - | Ductal | 3 | T4dN2aM0 | IIIC | Triple negative |
| I18 | IBC PBMC | 56-60 | White or Caucasian | Hispanic or Latino | No, never | - | - | - | Ductal | 3 | T4dN3aM0 | IIIC | Triple negative |
| I19 | IBC PBMC | 56-60 | White or Caucasian | Not Hispanic or Latino | No, never | - | - | - | Ductal | 3 | T4dN2M1 | IV | Triple negative |
| I20 * | IBC PBMC | 66-70 | White or Caucasian | Not Hispanic or Latino | No, never | - | - | - | Ductal | 3 | T4dN3cM0 | IIIC | Triple negative |
| I22 | IBC PBMC | 36-40 | White or Caucasian | Not Hispanic or Latino | Yes, currently smoke | ++ | ++ | - | Ductal | 3 | T4dN1M0 | IIIB | Luminal A |
| I23 | IBC PBMC | 46-50 | White or Caucasian | Declined to Answer | No, never | ++ | ++ | - | Ductal | 3 | T4dN1M0 | IIIB | Luminal A |
| I24 | IBC PBMC | 71-75 | White or Caucasian | Not Hispanic or Latino | Former | ++ | ++ | - | Mixed Ductal and Lobular | 2 | T4dN1M0 | IIIA | Luminal A |
| I25 | IBC PBMC | 80-85 | White or Caucasian | Not Hispanic or Latino | No, never | ++ | ++ | - | Ductal | 3 | T4dN1M0 | IIIB | Luminal A |
| I26 | IBC PBMC | 66-70 | White or Caucasian | Not Hispanic or Latino | No, never | ++ | - | - | Ductal | 3 | T4dN3cM0 | IIIB | Luminal A |
| I27 | IBC PBMC | 36-40 | White or Caucasian | Not Hispanic or Latino | No, never | - | - | - | Ductal | 3 | T4dN1M0 | IIIC | Triple negative |
| I28 | IBC PBMC | 46-50 | White or Caucasian | Not Hispanic or Latino | No, never | - | - | - | Ductal | 3 | T4dN3M0 | IIIC | Triple negative |
| I29 | IBC PBMC | 36-40 | White or Caucasian | Not Hispanic or Latino | No, never | - | - | - | Ductal | 3 | T4dN3cM0 | IIIC | Triple negative |
| I30 | IBC PBMC | 46-50 | Other | Hispanic or Latino | No, never | - | - | - | Ductal | 3 | T4dN3cM0 | IIIC | Triple negative |
| I31 | IBC PBMC | 51-55 | White or Caucasian | Not Hispanic or Latino | No, never | ++ | - | - | Ductal | 3 | T4dN1M0 | IIIC | Luminal A |
| I32 | IBC PBMC | 56-60 | White or Caucasian | Hispanic or Latino | No, never | - | - | - | Ductal | 2 | T4dN1M0 | IIIC | Triple negative |
| I33 | IBC PBMC | 56-60 | White or Caucasian | Not Hispanic or Latino | Former | - | - | - | Ductal | Unknown | T4dN3aM0 | IIIB | Triple negative |
| I34 | IBC PBMC | 51-55 | White or Caucasian | Not Hispanic or Latino | Former | - | - | - | Ductal | 3 | T4dN3M0 | IIIC | Triple negative |

| | | | | | | | | | | | | | |
|---|---|---|---|---|---|---|---|---|---|---|---|---|---|
| I35 * | IBC PBMC | 46-50 | Black or African American | Not Hispanic or Latino | No, never | - | - | - | Ductal | 2 | T4dN3M0 | IIIC | Triple negative |
| N01 | non-IBC PBMC | 41-45 | White or Caucasian | Declined to Answer | No, never | ++ | ++ | - | Ductal | 3 | T3N3aM0 | IIIB | Luminal A |
| N02 | non-IBC PBMC | 76-80 | White or Caucasian | Not Hispanic or Latino | No, never | ++ | ++ | - | Ductal | 3 | T3N1M0 | IIIA | Luminal A |
| N03 | non-IBC PBMC | 56-60 | Black or African American | Not Hispanic or Latino | No, never | - | - | - | Ductal | 3 | T2N1M0 | IIIB | Triple negative |
| N04 | non-IBC PBMC | 41-45 | White or Caucasian | Not Hispanic or Latino | Former | ++ | ++ | - | Ductal | 2 | T4bN1M0 | IIIB | Luminal A |
| N05 | non-IBC PBMC | 36-40 | Asian | Not Hispanic or Latino | No, never | - | - | - | Ductal | 3 | T2N1M0 | IIIB | Triple negative |
| N06 | non-IBC PBMC | 51-55 | White or Caucasian | Not Hispanic or Latino | No, never | - | - | - | Ductal | 3 | T2N1M0 | IIIB | Triple negative |
| N07 | non-IBC PBMC | 56-60 | Black or African American | Not Hispanic or Latino | Former | - | - | - | Ductal | 3 | T2N1M0 | IIIB | Triple negative |
| N08 | non-IBC PBMC | 51-55 | Other | Hispanic or Latino | No, never | - | - | - | Ductal | 2 | T3N1M0 | IIIB | Triple negative |
| N09 | non-IBC PBMC | 61-65 | White or Caucasian | Hispanic or Latino | No, never | ++ | - | - | Ductal | 3 | T2N3aM0 | IIIC | Luminal A |
| N10 | non-IBC PBMC | 36-40 | White or Caucasian | Not Hispanic or Latino | No, never | ++ | ++ | - | Ductal | 3 | T4bN3aM0 | IIIB | Luminal A |
| N11 | non-IBC PBMC | 51-55 | White or Caucasian | Not Hispanic or Latino | No, never | - | - | - | Ductal | 3 | T3N0M0 | IIIB | Triple negative |
| N12 | non-IBC PBMC | 66-70 | Black or African American | Not Hispanic or Latino | No, never | - | - | - | Ductal | 2 | T4bN1M0 | IIIC | Triple negative |
| N13 | non-IBC PBMC | 76-80 | Asian | Not Hispanic or Latino | No, never | - | - | - | Ductal | 3 | T2N1M0 | IIIB | Triple negative |
| N14 | non-IBC PBMC | 46-50 | White or Caucasian | Hispanic or Latino | Former | - | - | - | Ductal | 3 | T2N1M0 | IIIB | Triple negative |
| N15 | non-IBC PBMC | 51-55 | White or Caucasian | Not Hispanic or Latino | Former | - | - | - | Ductal | 2 | T3N2M0 | IIIB | Triple negative |
| N16 | non-IBC PBMC | 51-55 | Other | Hispanic or Latino | No, never | ++ | ++ | - | Ductal | 3 | T3N2aM0 | IIIC | Luminal A |
| N17 | non-IBC PBMC | 51-55 | Black or African American | Not Hispanic or Latino | No, never | ++ | - | - | Ductal | 3 | T3N1M0 | IIIB | Luminal A |
| N18 | non-IBC PBMC | 66-70 | White or Caucasian | Not Hispanic or Latino | No, never | ++ | ++ | - | Ductal | 2 | T2N3aM0 | IIIA | Luminal A |
| N19 | non-IBC PBMC | 61-65 | White or Caucasian | Hispanic or Latino | No, never | ++ | ++ | - | Ductal | 3 | T3N1M0 | IIIA | Luminal A |
| N20 | non-IBC PBMC | 31-35 | White or Caucasian | Hispanic or Latino | No, never | - | - | - | Ductal | 3 | T3N3bM0 | IIIC | Triple negative |
| N21 | non-IBC PBMC | 66-70 | White or Caucasian | Hispanic or Latino | Former | ++ | ++ | - | Ductal | 3 | T4bN2M0 | IIIB | Luminal A |

| | | | | | | | | | | | | | |
|---|---|---|---|---|---|---|---|---|---|---|---|---|---|
| N22 | non-IBC PBMC | 46-50 | White or Caucasian | Hispanic or Latino | No, never | - | - | - | Ductal | 3 | T3N0M0 | IIIB | Triple negative |
| N23 | non-IBC PBMC | 56-60 | White or Caucasian | Hispanic or Latino | Former | - | - | + | Ductal | 3 | T3N0M0 | IIIB | HER2+ |
| N24 | non-IBC PBMC | 26-30 | White or Caucasian | Hispanic or Latino | No, never | ++ | - | - | Ductal | 3 | T3N2aM0 | IIIB | Luminal A |
| N25 | non-IBC PBMC | 66-70 | White or Caucasian | Not Hispanic or Latino | No, never | - | - | - | Ductal | 2 | T2N1M0 | IIIB | Triple negative |
| N26 | non-IBC PBMC | 76-80 | White or Caucasian | Not Hispanic or Latino | Former | ++ | ++ | - | Ductal | 3 | T4bN0M0 | IIIB | Luminal A |
| N27 | non-IBC PBMC | 51-55 | White or Caucasian | Not Hispanic or Latino | Former | ++ | + | - | Ductal | 3 | T2N1M0 | IIIA | Luminal A |
| N28 | non-IBC PBMC | 56-60 | White or Caucasian | Not Hispanic or Latino | No, never | ++ | ++ | - | Ductal | 3 | T2N3M0 | IIIA | Luminal A |
| N29 | non-IBC PBMC | 31-35 | Other | Hispanic or Latino | No, never | - | - | - | Ductal | 3 | T2N1M0 | IIIB | Triple negative |
| N30 | non-IBC PBMC | 41-45 | Black or African American | Not Hispanic or Latino | No, never | ++ | ++ | - | Lobular | 3 | T3N1M0 | IIIA | Luminal A |
| N31 | non-IBC PBMC | 66-70 | White or Caucasian | Not Hispanic or Latino | No, never | - | - | - | Ductal | 2 | T2N1M0 | IIIB | Triple negative |
| N39 | non-IBC PBMC | 36-40 | Asian | Not Hispanic or Latino | No, never | - | - | - | Ductal | 3 | T3N0M0 | IIIB | Triple negative |

* These patients received treatment before sample collection.

Donor IDs are notated as: H, Healthy; N, non-IBC; I, IBC.
Age and Smoking conditions are based on the time of sample collection.
ER+/- and PR+/- categories use the same classification scaling:

- : 0 to less than 1%
+ : 1 to less than 10%
++ : greater than or equal to 10%
Both + and ++ were considered positive for subsequent Subtype classification.

Tumor, Node, Metastasis (TNM) staging system (www.mdanderson.org).
Tumor subtypes are based on ER/PR/HER2 expression:

Luminal A : either ER+ or PR+, and HER2-
Luminal B: either ER+ or PR+, and HER2+
HER2+: ER- and PR- and HER2+
Triple negative: ER- and PR- and HER2-

NA: Not Available.
